# How do clinicians conceptualise negative self-concept in complex post-traumatic stress disorder (CPTSD)?

**DOI:** 10.1101/2025.06.09.25329248

**Authors:** Adele Melia, Nicola Morant, Hana Afrah, Sasha Menon, Cornelius Katona

## Abstract

Negative self-concept (NSC) is one of the core ‘disturbances in self-organisation’ (DSO) symptoms of CPTSD. It is characterised by persistent negative beliefs about the self and is common in traumatised populations who have endured crises such as childhood sexual abuse survivors, refugee groups, and military veterans. However, there is limited research on how NSC is conceptualised and managed in CPTSD and other related disorders. This qualitative study explores how clinicians conceptualise NSC in these populations. Semi-structured interviews were conducted with 15 clinicians (clinical psychologists, psychiatrists and psychotherapists) and data was analysed using thematic analysis. Clinicians described NSC as involving a persistent negative view encompassing feelings of self-blame, shame, guilt, and worthlessness. Early childhood attachments and systemic responses following traumatic experiences were identified as influential in shaping self-concept. Facilitators to healing included self-compassion, the therapeutic relationship and social re-integration. These findings aim to offer insights for clinicians supporting traumatised populations.

## 1. Introduction

The International Classification of Diseases, 11th ed. (ICD-11; World Health Organisation, 2019) included a new diagnosis of complex post-traumatic stress disorder (CPTSD). CPTSD is more likely than PTSD to result from exposure to multiple and prolonged traumatic crises in which escape is difficult or impossible (Brewin et al., 2017). PTSD is characterised by three symptom clusters: re-experiencing the trauma in the present, avoidance of traumatic reminders and a persistent sense of threat. CPTSD requires the presence of three additional domains of ‘Disturbances in Self-Organisation’ (DSO): affect dysregulation, difficulties in relationships, and Negative Self-Concept (NSC). NSC is characterised by persistent negative beliefs about the self, which has been ’defeated’ by trauma (ICD-11, 2019).

Whilst most research on CTPSD focuses on survivors of childhood sexual abuse (Cloitre et al., 2019; Palic et al., 2016), CPTSD is also common in other traumatised populations, including refugees and asylum seekers (de Silva et al., 2021; Nickerson et al., 2016) and military veterans (Murphy et al., 2020). Whilst multiple traumatic events in adulthood are associated with both CPTSD and PTSD, childhood trauma may be a particular risk factor for CPTSD (Cloitre et al., 2019; Van der Kolk et al., 2005). Military veterans with CPTSD show higher rates of childhood adversity than those with PTSD (Murphy et al., 2021).

However, Palic et al., (2016) found that risk of CPTSD varied according to the nature of the trauma, regardless of the developmental stage at which it is experienced. Survivors of childhood sexual abuse and refugees and asylum seekers were both more likely to develop CPTSD than military veterans. The authors suggested that the more ‘interpersonal’ (i.e caused by the cruelty of others) the trauma, the greater the likelihood of developing DSO symptoms. Interpersonal trauma may generate persistent feelings of shame and guilt leading to NSC in CPTSD (Dorahy et al., 2017).

CPTSD has a prevalence of 16-38% in refugees and asylum seekers who have often experienced multiple interpersonal traumas, including trafficking and exploitation, in their country of origin and during their flight (de Silva et al., 2021). Additionally, they may face post-migration difficulties and lack of social support which have also been associated with DSO symptoms (Hecker et al., 2018). Uncertain legal status, difficult living conditions, discrimination and difficulty integrating into communities may perpetuate NSC.

More treatment-seeking military veterans fit criteria for CPTSD than for PTSD (Murphy et al., 2020). Those with CPTSD report higher rates of ‘moral injury’ (emotional and cognitive responses following a moral transgression) than those with PTSD (Currier et al., 2021). Shame, guilt and anger are complex emotions associated with moral injury and closely relate to NSC, suggesting that the meaning attached to these traumatic experiences rather than their type may increase risk of CPTSD.

Network analyses of CPTSD symptoms have found NSC to be its most central feature (Knefel et al., 2016; Knefel et al., 2019), suggesting that targeting NSC may have a positive cascade effect on other symptoms (Karatzias et al., 2019; McNally et al., 2015). However, the evidence base for treating NSC in CPTSD is limited. A meta-analysis by Banz et al., (2022) concluded that, in people with PTSD, psychological interventions significantly improve NSC, yet there is no consensus on what treatments work best for NSC or what factors determine a particular intervention’s effects. To our knowledge, no study has looked specifically at treatment of NSC in CPTSD. People with CPTSD may need treatment for longer than those with PTSD or may require a variety of treatment strategies, especially for DSO symptoms (Cloitre et al., 2019). Using phase-based interventions, which involve stabilisation to develop safety before trauma memory processing, has benefited individuals with CPTSD (Cloitre et al., 2011), but this may delay access to effective trauma-focussed treatment (de Jongh et al., 2016).

NSC is also a central characteristic of other disorders. Negative beliefs about oneself and the world, with heightened levels of self-blame and worthlessness are distinctive features of depression (Harrison et al., 2022). Negative self-appraisal is also a risk factor for PTSD (O’Donnell et al., 2007). This raises the question of whether NSC is conceptualised similarly between disorders or has distinct features within CPTSD.

Given the lack of consensus and evidence regarding the CPTSD construct and optimal therapeutic strategies, understanding the role and conceptualisation of NSC within CPTSD may help distinguish CPTSD from other disorders and inform treatment. Exploring how clinicians expert in working with complex trauma understand and treat NSC and how they think it differs between populations may inform evidence-based management and treatment plans. This study uses qualitative methods that allow in-depth explorations of complexities. We focus on clinicians’ experiences of working with NSC in three key CPTSD groups: childhood sexual abuse survivors, refugees and asylum seekers, and military veterans.

## 2. Methods

Methods are reported in line with COREQ guidelines (Tong et al., 2007). The study was approved by the University’s ethics committee on 10/03/2021. All participants provided written informed consent prior to participating and for their anonymised data to be published.

### 2.1 Participants

Participating clinicians were from London-based community mental health services, including charities and the National Health Service. Services either specialised in trauma or accepted traumatized patients. Recruitment began on 10.03.2021 and ended on 10.03.2023. Participants were identified initially through professional networks and then approached via email. A copy of the recruitment email template has been provided in **Supplementary Materials.** Snowball sampling was used to recruit further participants to ensure diversity including clinicians working across trauma populations (childhood sexual abuse survivors, refugees and asylum seekers, and military veterans) and from various professional backgrounds (psychologists, psychiatrists and psychotherapists).

### 2.2 Procedures

The interview topic guide was developed by a research team including a senior psychiatrist working with trauma populations (CK) and a qualitative mental health researcher (NM); a copy of this topic guide can be found in **Supplementary Materials**. Questions covered participants’ understandings of the diagnosis of CPTSD, NSC, comparisons across disorders, and treatment strategies for the population they work with. Semi-structured interviews were conducted by HA and AM, both postgraduate students and aspiring clinical psychologists with interests in trauma.

Interviews were conducted via telephone or video call and lasted 30-60 minutes. Interviewers recorded initial impressions and a reflexive summary of each interview. Interview recordings were transcribed verbatim (by HA and AM) with identifying information removed and pseudonyms assigned to interviewees.

### 2.3 Data Analysis

Data was analysed using thematic analysis (Braun & Clark 2006) within NVivo 12 software. Analysis combined inductive and deductive approaches to explore participants’ experiences and views whilst developing relevant themes. The six phases of thematic analysis described by Braun & Clark (2006), were used recursively. A researcher (AM) familiarised herself with the interview recordings and transcripts. Codes were developed to reflect the meaning constructed by participants and the researcher’s reflexive interpretation, then explored and organised into a hierarchical structure of themes and sub-themes. Themes were further refined and named to reflect the dataset and research questions. To enhance analytic validity, AM made reflexive notes on judgements, reasoning and emotional reactions throughout the analytic process, and held regular meetings with CK and NM to discuss developing themes, introduce different perspectives, and maintain reflexivity.

## 3. Results

Fifteen clinicians were interviewed (Table 1). Some practiced in multiple roles and with different trauma populations. Their patients were survivors of childhood trauma, refugees/asylum seekers, and military veterans. Most participants were female, white British, and aged 30-40. Psychologists employed various theoretical orientations, psychiatrists used predominantly medicalised approaches, whilst psychotherapists were psychoanalytically informed. One interviewee experienced technical difficulties and was only able to answer half the Topic Guide questions.

**Table 1.** Sample Characteristics.

|  |  |
| --- | --- |
| Sex |  |
| Female | 12 |
| Male | 3 |
| Age |  |
| 30-40 | 7 |
| 40-50 | 5 |
| 50+ | 3 |
| Ethnicity |  |
| White British | 11 |
| White other | 1 |
| South Asian | 1 |
| Chinese | 1 |
| British Asian | 1 |
| Professional Background |  |
| Clinical Psychologist | 10 |
| Psychiatrist | 4 |
| Psychotherapist | 2 |
| Trauma Population |  |
| Childhood trauma/sexual abuse | 12 |
| Refugees and Asylum Seekers | 11 |
| Military Veterans | 4 |
| Theoretical Orientation/Treatment Approaches |  |
| CBT | 6 |
| EMDR | 6 |
| CFT | 4 |
| MBT | 1 |
| ACT | 2 |
| CT-PTSD | 2 |
| NET | 6 |
| Dynamic | 2 |
| TF-CBT | 1 |
| Trauma informed | 2 |
| Medical | 4 |
| Psycho-analytically informed | 2 |

*Note. ACT = Acceptance and Commitment Therapy; CBT = Cognitive Behavioural Therapy; CT-PTSD = Cognitive Therapy for Post-traumatic Stress Disorder; CFT= Compassion Focussed Therapy; EMDR = Eye Movement Desensitisation and Reprocessing; MBT = Mentalisation Based Therapy; NET = Narrative Exposure Therapy*

Analysis produced four main themes reflecting clinicians’ perspectives on NSC in CPTSD and across trauma populations. They discussed vulnerability and protective factors for NSC, specifically the impact of early attachments and the presence or absence of a whole systems approach. They explored treatment strategies including the importance of treating NSC, as well as the benefits of self-compassion, the therapeutic relationship and societal re-integration. Finally, participants discussed comparisons of NSC across mental health disorders.

### 3.1 How clinicians understand NSC

It was evident that NSC was regarded as a core aspect of CPTSD. Clinicians described it as involving a persistent and profound negative view often encompassing feelings of self-blame, shame, guilt, and worthlessness. These beliefs were viewed as deeply rooted in an individual’s sense of self. Most clinicians considered NSC to be especially difficult to shift when working with patients - often because they incorporate emotions that are complex and difficult to explain:

> Sue: *“I think if there’s just very high levels of shame and guilt um, these are very, very difficult emotions for people to want to open up cause they’re very painful but also… it can take quite a lot of time to really understand where did that belief come from.”*

#### 3.1.1 Self-Blame Across Trauma Populations

Self-blame was a prominent feature identified by clinicians working across different trauma populations. The multiplicity of traumatic events experienced led patients to believe they must be to blame for their trauma and do not *‘deserve to have good things’* (Rose). However, some clinicians reported differences in its manifestation across trauma populations.

Generally, clinicians felt that patients who had experienced childhood trauma, sexual abuse, and trafficked refugees and asylum seekers, often asked ’why me?’. It appeared difficult for such patients to understand why the trauma had happened specifically to them, consequently feeling that they must have done *‘something to deserve this’* (Debbie). The interpersonal nature of this trauma complicates emotions further because there is a sense that it is only happening to them, therefore there is something wrong with them.

> Joe: *“I can’t think of any people who experienced that, especially kind of repeated interpersonal trauma, uhm, where there wasn’t some level of self-blame or some level of thinking about, why is this happening to me?”*

For refugees and asylum seekers, some clinicians felt that self-blame sometimes manifested because they were unable to protect others:

> *Alice: “I was just thinking about an Albanian young person, and he sort of has this profound belief that he should have protected, particularly his sister, from traffickers.”*

Similarly, clinicians working with military veterans expressed that experiencing moral injury during a traumatic event often complicates how their patients appraise the traumatic event and where they place blame. If they engage in or fail to prevent an act that goes against their moral values or feel betrayed by others, this contributes to their NSC.

Conversely, some clinicians described how refugees and asylum seekers might be able to attribute meaning to their traumatic experiences that protects them from self-blame. For instance, understanding trauma experiences in the context of political activism may be protective if they continue to believe that they are achieving something:

> Fatima: *“I think that there is, there is certainly that feeling of being able to attribute some meaning to what has happened, and then being part of something greater than merely yourself.”*

#### 3.1.2 Self-blame, Shame, Guilt and Worthlessness are interlinked

Across populations, clinicians described patients having high levels of shame and guilt, often linked to the trauma they experienced. These beliefs are perpetuated by perceptions of how others view them - for example that *’other people are going to think negatively of me*’ (Debbie) and ‘*they’ll reject me’* (Sue). This may generate feelings of guilt that they should have done something differently to prevent the trauma. These feelings appear interlinked with how patients view themselves. For instance, self-blame, shame and guilt maintain patients’ feelings of worthlessness and being undeserving. This is likely to persist due to the multiple traumatic events they experienced.

### 3.2 Vulnerability and Protective Factors for Negative Self-Concept

#### 3.2.1 Early Attachments are key

Lack of early attachments and high rates of childhood trauma in those with CPTSD was a common theme. Most clinicians perceived CPTSD (and NSC in particular) to result from childhood trauma, which contributed to their patients having such strongly held and persistent negative beliefs (i.e. their NSC) about themselves.

Some clinicians however considered that sufficiently severe multiple traumatic events (such as torture and trafficking) in adulthood can cause CPTSD even in the absence of childhood trauma. However, patients traumatised in adult life but with positive attachments and relationships early in life may find it easier to work through their negative self-beliefs. Across trauma populations, lack of positive childhood attachments and childhood trauma were seen as risk factors for CPTSD. Lack of positive early attachments was deemed more of a risk factor for CPTSD than for PTSD:

> Alice: *“who do you end up giving a diagnosis of, you know PTSD compared to complex PTSD… although they may have had very similar experiences say in their journey to Libya… they don’t end up coming out with complex PTSD, it’s PTSD. And I think, you know, a lot has to do with having very good early attachment relationships”*.

Clinicians explained that lack of positive attachments in early childhood may lead to NSC because it shapes how individuals conceptualise their sense of self and is a foundation for later experiences. Subsequent traumatic events in adulthood feed into their feelings of worthlessness and notions that they are to blame for their traumas.

> Kate: *“So you can’t shift it ’cause they always feel it, they didn’t just feel it during the trauma, they felt it before the trauma as well, very much so. So those are the-that’s the subgroup of people that, that I would say have the negative self-concept bit of complex PTSD.”*

#### 3.2.2 A Whole Systems Approach is Required

Many clinicians explained how the system around an individual after a traumatic event can affect how they make sense of the event and, in turn, their sense of self. For children who have been abused, clinicians discussed the importance of how the system responds to the child and the care they receive. Family members, as well as service providers, influence how the child appraises the traumatic event and how their self-concept is subsequently influenced.

> Candice: *“if people are protective and believing and uhm and supportive and comforting and really helps the child normalise and understand their reaction to what’s happened as well, then they can really be protected from the more negative aftereffects.”*

Similarly, clinicians working with military veterans believed that a supportive environment within military culture may be protective. One clinician felt that veterans’ post-traumatic growth and the system around them may be protective against profound negative feelings and beliefs attached to the traumatic event. Conversely, an unsupportive system was considered detrimental to self-concept. This was most evident for refugees and asylum seeker populations who are often faced with further challenges when they arrive in their host country, including cultural barriers and difficulties accessing funding, housing and employment.

> Poppy: *“the process that they go through here is so abusive and so invasive and so dehumanising that for clients who at the beginning of their process perhaps had a straightforward sense of trauma and felt about themselves, uhm as someone trying to overcome something. Two, three, five, ten years later, the system has really added a layer of complexity to their trauma that perhaps wasn’t there to begin with.”*

For clinicians working with refugees and asylum seekers, it was evident external factors perpetuating NSC were one of the most difficult aspects complicating treatment. Many clinicians considered that a whole-systems approach in which services work collaboratively was necessary for individuals with CPTSD to gain stability across different aspects of their lives and progress in treatment. However, this was seen as often being out of clinicians’ control:

> Sara: “*Yeah, but then, I can’t sort out the fact that, actually, they’re experiencing financial difficulties as a result of, maybe, the failure of other services to support them, or maybe government cuts and how they’ve impacted on them, or maybe the fact that, actually, they’re living in an area, where, actually, they’re being targeted or oppressed. Or the fact that actually, the family is experiencing racism from, you know, particular, um, organisation, you know, local authorities, education and so forth.”*

### 3.3 Treatment Strategies

#### 3.3.1 Treating Negative Self-concept is Essential

NSC was seen as a potential barrier that often complicated the treatment process. Clinicians discussed how therapy typically takes longer for patients with CPTSD, compared to those with PTSD, because their NSC beliefs are difficult to shift. Most clinicians believed that treating feelings of shame, guilt and self-blame, whilst tailoring treatment to the individual, was essential. There was some divergence about whether to treat symptoms of NSC before or after going into trauma-focussed work. Many clinicians thought that treating NSC first was vital because these beliefs are likely to cause considerable distress, and it was important not to re-shame their patients:

> Eve: *“But I think there’s probably a lot more work to be done on the negative kind of appraisals and self-concept before you can go into reliving with the complex PTSD end of the scale…otherwise, you’re just going to re-shame them by doing the re-experiencing.”*

Others thought these beliefs could be addressed during trauma-focussed work and that treating core PTSD symptoms would itself have a positive effect on NSC.

> Sue: *“it’s a bit more of an implicit like um approach, hoping that the negative concept will improve as the trauma memory is processed and then strengthening positive self-beliefs as they come.”*

Two participants thought that trauma-focused work should come first because addressing unprocessed memories would help patients to function better in everyday life. Reducing core symptoms of PTSD was also deemed necessary to give patients the headspace to understand and work on negative trauma-related beliefs.

#### 3.3.2 Self-Compassion

Many clinicians thought that developing self-compassion was a powerful way to shift negative beliefs. Normalising how patients feel helped ‘de-shame’ some of their beliefs about themselves and opened possibilities that the trauma was not their fault. Some participants mentioned soothing techniques used in Compassion Focussed Therapy (CFT) to relieve physiological responses to trauma, such as breathing and cultivating a safe space. Participants whose theoretical background was in CFT discussed how it can alleviate shame and self-blame beliefs and is therefore useful for treating NSC. Others thought that using CFT was useful in conjunction with other types of therapies:

> Joe: *“I had experiences with people in EMDR where you might hammer away for session after session, and then get blocked over some belief. It was a negative self-belief, uhm, but then introducing CFT into EMDR changed the perspective.”*

### 3.4 Therapeutic Relationship

Almost all participants mentioned that the therapeutic relationship was a key factor in treating NSC in CPTSD. Many of their patients had suffered a breakdown of relationships in the context of their traumatic experiences. The therapeutic relationship was an opportunity for clinicians to help their patients re-build trusting relationships with others.

> *Rose: “the complex PTSD presentation is often, more often than not, the result of like damaging interpersonal relationships, damaging interpersonal trauma, and a healing therapeutic relationship can be very powerful in my experience.”*

Some clinicians brought discussions of the therapeutic relationship back to attachments and early relationships. Their patients found it more difficult to build a trusting therapeutic relationship if their early attachment relationships had been shattered. This has implications for treatment and often makes the therapy process longer as patients’ ability to trust needs to be re-built. Creating a strong therapeutic alliance in these circumstances was therefore deemed particularly important by clinicians.

> Lucy: *“I think any treatment that gives someone the opportunity to be able to build an attachment relationship and work through some of this is helpful”*

The therapeutic relationship was also seen as creating a space within which patients felt believed and validated. This empowered them to feel valued in society. Through building a therapeutic relationship, clinicians were able to build trust and facilitate disclosure of traumatic experiences. Once trust had been established, it was easier to test out patients’ shame-based beliefs, which then helped them to resolve their NSC.

### 3.5 Re-integration

Some clinicians felt strongly that helping patients re-integrate into society was important as well as indicating that treatment was effective. Mentioned examples included getting a job, volunteering, doing well in education, sustaining positive relationships, and taking up a hobby. Finding a purpose was seen as evidence of improvement in NSC. Progress in re-integration often helped patients feel better about themselves, with a sense that they had something positive to contribute.

> Stephen: *“And then once they start to do things their sense of self-worth improves, like I say not because of the therapy, but it’s increased because of, where we all get our self-worth from, you know you start doing things and getting meaning.”*

### 3.6 Comparisons with other Mental Health Disorders

Almost every participant highlighted common mental health comorbidities in CPTSD, including PTSD, bipolar disorder, depression and anxiety. Clinicians found it challenging to tease out distinct aspects of disorders due to the overlap of symptoms. When reflecting specifically on the qualitative differences of NSC in CPTSD compared to other disorders, clinicians found this difficult to clarify:

> Sue: *“like where does complex PTSD and the negative self-concept and all that, and all the impact that that has on a person…where does that end, and depression begin.”*

Some clinicians saw no benefit in discerning differences in NSC across diagnoses, with NSC perceived as a common symptom across many disorders. Symptom overlap may however make it difficult to determine which disorder best describes a patient’s condition and may complicate treatment because it generates a *‘world of chaos with different clinicians doing different things’* (Stephen). However, many clinicians recognised the added value of formulation in understanding NSC for each individual and approaching it from a trauma perspective, including its origins and persistence.

The most apparent difference when comparing NSC in CPTSD and other disorders was its profundity in CTPSD. Some clinicians observed that negative self-beliefs in CPTSD are more difficult to shift due to their deep-rooted nature, unlike PTSD where negative beliefs may stem from peritraumatic distress:

> Debbie: *“And I guess often tied to trauma memories, uhm, but I suppose in a different way to how it might be with PTSD, where it might be linked to the memories. So, I feel bad, or guilt or shame, but about this particular memory. Complex PTSD, I tend to see it as the shame and guilt is about myself as a whole person. Uhm, rather than only about the memory.”*

Similarly, NSC in depression may be a function of the depressive episode and therefore fluctuate. In CPTSD, however, NSC is more persistent, reflecting the multiple traumatic events someone experiences, which often begin during childhood.

> Alice: “*But for me, there’s something about the complex PTSD, the negative self-concept, it’s that much more consistent, sustained sort of presence. Whereas depressive disorder, you may have a history where you know actually, they were functioning better, they didn’t have those thoughts that this is very much linked to the depressive episode.”*

The prolonged and profound nature of NSC in CPTSD was deemed more difficult to treat than other disorders.

## 4. Discussion

This study provides important insights into how clinicians working with different trauma populations conceptualise NSC and explores their views on effective ways of addressing it. Clinicians generally view NSC as a profound negative view of oneself encompassing feelings of self-blame, shame, guilt and worthlessness. Despite commonalities in how clinicians conceptualise NSC across trauma populations, differences emerged, particularly regarding self-blame. For individuals experiencing childhood trauma or trafficking, clinicians attributed self-blame to the interpersonal nature of this trauma and survivors’ consequent struggle to comprehend why they suffered such experiences. Refugees and asylum seekers may also blame themselves if they are unable to protect loved ones (Donovan et al., 2025) but may be able to attribute more meaning to their experiences. The further difficulties they face following their arrival in their host country adds complexity to their trauma. DSO symptoms have previously been linked to post-migration difficulties (Hecker et al., 2018; Nowak et al., 2023). NSC may be perpetuated by these difficulties. Military veterans may experience moral injury during a traumatic event, which complicates how they subsequently construct meanings of their experience and in turn contributes to their NSC. Military veterans with CPTSD are particularly vulnerable to moral injury and associated distress (Currier et al., 2021).

Clinicians indicated that childhood trauma is a consistent risk factor for developing CPTSD, regardless of the trauma population. This is supported by research findings in military veterans (Murphy et al., 2021) and in refugees and asylum seekers (Schiess-Jokanovic et al., 2021). Clinicians’ views align with developmental theory, which suggests that early exposure to interpersonal trauma and disruptions in caregiving impact neural development and attachment (van der Kolk, 2005). This influences how individuals subsequently view themselves and the world because of their tendency to incorporate negative beliefs into their self-concept. However, this interaction is complex and not necessarily linear. Some research suggests that those exposed to childhood adversity are more likely to join the military to escape dysfunctional home environments (Katon et al., 2015). Childhood trauma is also a key risk factor for trafficking victimisation (Choi, 2015). The ICD-11 diagnosis of CPTSD does not require a history either of multiple trauma or of childhood trauma. Some individuals meet ICD-11 CPTSD criteria having only experienced a single traumatic incident in adulthood (Cloitre et al., 2019).

We also explored clinicians’ views of NSC in CPTSD compared to other disorders. Clinicians were tentative when discussing differences. However, not all clinicians were asked explicitly about potential similarities or differences, so these results were analysed from a smaller sample and should be viewed with more caution. Exploring these differences in future research may be necessary for informing treatment for CPTSD. A few clinicians considered NSC to be more prolonged in CPTSD than in other disorders, therefore requiring longer-term treatment.

### 4.1 Clinical and Research Implications

This study highlights differences in how NSC manifests in different populations and highlights some important strategies for treatment. There was a consensus across clinicians that negative beliefs about the self were difficult to shift and often a barrier to treatment. Clinicians in the current study often adopted a compassionate framework in therapy to target negative beliefs about the self and believed it to be beneficial. However, evidence for the effectiveness of compassion-focussed interventions for CPTSD remains limited (Karatzias et al., 2019).

The therapeutic alliance was regarded as essential for helping patients with CPTSD, specifically in supporting patients to share their negative beliefs about themselves. A recent systematic review investigating therapeutic alliance in PTSD found it to be a significant predictor for outcomes, over and above the type of therapy provided (Howard et al., 2022). Given the additional DSO symptoms in CPTSD, building a strong therapeutic alliance becomes even more critical to support these individuals. Optimising clinician training and support in establishing a positive therapeutic relationship with CPTSD patients is essential.

The ongoing debate regarding a phase-based approach to CPTSD treatment was reflected in this study. In a survey of 50 trauma experts, 84% endorsed a phase-based approach to treating CPTSD (Cloitre et al., 2011). Our findings align with this, with most clinicians emphasising the need to address negative self-beliefs before engaging in trauma-focused interventions, particularly to prevent re-shaming their patients. Additionally, clinicians highlighted the importance of the re-integration phase for improving NSC. This phase has been given little attention in research (Purnell et al., 2021), and warrants further exploration.

Future research should examine protective and vulnerability factors for NSC and CPTSD across trauma populations. This will aid in understanding the causes of CPTSD, identifying at-risk individuals and determining the need for extended and/or specific treatment. Examining the role of attachment in the development of DSO symptoms and CPTSD may be useful. Preliminary results suggest a relationship between fearful attachment styles and CPTSD symptoms (Karatzias et al., 2021). Further research could also explore the necessity for a phase-based approach, the importance of re-integration for NSC, and the mechanisms by which compassion-focussed strategies resolve NSC.

### 4.2 Strengths and Limitations

A strength of the current study is the inclusion of clinicians from diverse professional backgrounds working with various trauma populations. This helped gain a broad understanding of how NSC is conceptualised and worked with in CPTSD treatments across trauma populations.

Nonetheless, there were several limitations. The sample consisted mainly of clinical psychologists with only two psychotherapists in the sample, and we only included two clinicians working with military veterans. Purposive and snowball sampling may have resulted in participants being predominantly from the same or similar organisations and may therefore reflect specific organisational values. Many of the clinicians interviewed had a research background, which may have shaped their accounts. This limitation might extend to a lack of inclusion of clinicians who practice Cognitive Processing Therapy (CPT), despite its widespread use in addressing NSC emotions such as inappropriate self-blame and post-traumatisation guilt (Stayton et al., 2018). Additionally, clinicians’ views are constructed through experience with treatment-seeking individuals, who may differ from those unable or unwilling to seek treatment. Finally, our study only allowed relatively limited exploration of how NSC manifested in disorders other than CPTSD. This suggests a potential avenue for future research, possibly requiring a different methodological approach.

## 5. Conclusion

This study highlights that clinicians consider NSC to be a core aspect of CPTSD that encompasses complex emotions and presents therapeutic challenges. These emotions are likely to manifest differently across trauma populations, which may be important when tailoring interventions for these individuals. Similarly, protective and vulnerability factors may be different depending on the trauma an individual experiences. However, participants agreed that childhood trauma is likely to be a significant predictor of NSC. Effective treatment for NSC remains uncertain, highlighting the need for further research to establish treatment guidelines for clinicians.

## Data Availability

The participants of this study gave written consent for their anonymised data to be published, however due to risks of deanonymisation, raw data is not available. Other materials such as the interview topic guide, and example recruitment email text has been provided in Supplementary Materials.

