## Supplemental materials for "How do clinicians conceptualise negative self-concept in complex post-traumatic stress disorder (CPTSD)?"

**Supplementary materials 1:** *Recruitment email sent to potential participants and their networks*

‘’Dear Sir or Madam,

I am writing to you about a research project that is being carried out for an MSc Dissertation as part of the Clinical Mental Health Sciences course at UCL. The title of the project is ‘how do clinicians conceptualise negative self-concept in people with complex PTSD (cPTSD) and in other mental health conditions?’

This dissertation is supervised by Prof Cornelius Katona ( – Consultant Psychiatrist at Helen Bamber Foundation & Honorary Professor at

UCL) and Dr Nicola Morant ( – Associate Professor at UCL).

You have been identified as a clinician who deals with people who have endured trauma and have a diagnosis/suspected diagnosis of Complex Post-Traumatic Stress Disorder (CPTSD) based on the ICD-11. We would like you to consider participating in this study.

We are interested in collecting some data from clinicians who work directly with trauma

populations. This qualitative study will be exploring how clinicians conceptualise negative self-concept in people with CPTSD and in other mental health conditions, using semi-structured interviews. Negative self-concept will be explored as a component of CPTSD, exploring how this presents in different populations and in different co-morbid disorders. It is hoped that this study will increase understanding of negative self-concept and potential treatment options due to the current limited literature on this in relation to CPTSD.

The study should take no more than an hour and can be in the format that best suits you (video call via MS Teams or telephone). We anticipate collecting data over the next two months.

I’d be grateful to hear back from you at about whether you might be interested to participate. I have attached the information sheet and consent form if you would like to read more about the study. The study has already received approval from UCL ethics committee (number is 18391/001) Please let me know if you have any questions or concerns and if you would like to take part in this study.

I look forward to hearing back from you. Thank you very much in advance.

Best wishes’’

**Supplementary materials 2**: *Draft interview topic guide*

**Introduction:**

In this study, we are interested in investigating negative self-concept, which is the view of a defeated sense of self that occurs from trauma and is characterised by persistent negative beliefs about self. We are particularly interested in how negative self-concept presents in people with complex PTSD and how this differs from other disorders and in different trauma populations. We are also interested in how these understandings can help in future treatment of this relatively new diagnosis. Therefore, the questions asked will focus on these topics. Thank you so much for volunteering your time and attention to take part in this study.

**Section 1: Introductory questions to gauge understanding**

The following questions will explore your general experiences and understanding of some of the concepts we will be exploring in subsequent sections.

- What is your understanding of the ICD-11 diagnosis of complex PTSD?

***Prompts****: disorders of self-organisation (DSO)? Negative self-concept in particular?*

- Do you work with people who are diagnosed with complex PTSD?

***Prompts:*** *how often? Is it a rarity or a common occurrence? Do you use any complex PTSD diagnostic measures (e.g., International Trauma Questionnaire) in your practice?*

- Which populations do you work with most regularly: refugees/asylum seekers, combat veterans or people who have experienced childhood sexual abuse?

***Prompts:*** *have you worked with one more than the others? Do you work with all three an even amount?*

**Section 2: Professional experiences with negative self-concept**

The following questions will have a more specific focus on your experiences with service users who present with negative self-concept in your line of work. These questions will require you to reflect on your clinical experiences with different clinical populations as well as differences between individuals. Feel free to give specific examples from prior experiences whilst answering questions.

- What is your experience of negative self-concept in people diagnosed with complex PTSD?

***Prompts****: what does it look like? What kinds of things do people who struggle with this typically say? What do they do? How does this compare with those who don’t struggle with negative self-concept?*

- What do you think is unique about the presentation of negative self-concept in the population(s) you work with?

***For clinicians who have worked with more than one of the clinical populations:*** *how has it differed between the populations? If not, has the presentation linked to their status in any way (e.g., linked to being a refugee/veteran/abuse victim)?*

- What differences have you found in the presentation of negative self-concept between individuals?

***Prompts****: nature of the trauma experience? Whether they experienced one or multiple traumas? Their current social situation (financial, settlement status, living environment etc)?*

**Section 4: Experiences with different disorders**

The following questions will require you to think more specifically about the types of comorbid mental health issues that the people you work alongside present with, as well as the differences between these disorders and complex PTSD.

- What are your experiences of negative self-concept in those with comorbid disorders alongside complex PTSD compared with those with a single diagnosis of complex PTSD?

***Prompts:*** *are there many people with only a mental health diagnosis of complex PTSD? Do the different disorders interact in any special way in presentation?*

- How do the presentations of negative self-concept in people with complex PTSD differ from people with depression?
- “ “ people with (non-complex) PTSD?
- “ “ people with any other disorders that come to mind?

**Section 5: Implications for treatment of negative self-concept**

This final question is about treatment implications.

- What do your experiences of people with negative self-concept tell you about how to treat this aspect in people with complex PTSD?

***Prompts:*** *what types of therapy may be beneficial?*

**Concluding questions:**

- Is there anything more you would like to share?
- Do you have any questions?
